# A modelling framework to improve antibody titer estimation from dilution series data: application to RSV Foci Reduction Neutralization Tests

**DOI:** 10.1101/2025.07.24.25332140

**Authors:** Yan Wang, Qianli Wang, Chris Wymant, Junyi Zou, Lan Yi, Meng Xu, James A. Hay, Hongjie Yu

## Abstract

Accurately determining antibody levels from experimental data is important for measuring population immunity, guiding vaccine development and informing public health policy. Based on raw dilution series data from respiratory syncytial virus (RSV) foci reduction neutralization tests (FRNTs), we identified notable bias and uncertainty in neutralizing antibody (nAb) titer estimates between batches when using standard, off-the-shelf methods such as the Karber formula and four-parameter logistic (4PL) model. To address these problems, we developed a Bayesian hierarchical modelling framework for estimating nAb titers, which corrects for batch effects and other sources of experimental variation. The model was shown in both simulations and real data from RSV serosurveys to provide more accurate and unbiased nAb titer estimates than existing approaches. Among all methods evaluated, the Karber formula exhibited the highest variability and led to increased rates of both false positives and false negatives. The 4PL model tended to produce negative estimates in samples with low antibody levels. These inaccurate individual-level estimates, in turn, biased population-level measures, including geometric mean titers (GMTs), seroprevalence, seroconversion rates and fold-rise rates. In contrast, the adjusted estimates from our model demonstrated the highest accuracy. Spearman correlation coefficients between estimated and true titers from simulated data were 0.63 for Karber estimates, 0.87 for 4PL estimates, and 0.96 from our model. This framework can be readily adapted to other antibody assays which produce dilution series data, and can enhance the accuracy and precision of titer estimates across a range of experimental settings.

## Introduction

Serological data are crucial for quantifying population exposure and immunity against a range of pathogens, particularly vaccine preventable diseases where humoral immunity provides a key mechanism of protection.^1–4^ By measuring pre-existing antibody levels induced by past infection and vaccination, public health interventions can be targeted to susceptible individuals and populations at greatest risk of disease. This approach to characterizing population immunity is particularly timely for respiratory syncytial virus (RSV) following recent advances in RSV prophylactics, including the approval of several vaccines and a half-life-extended monoclonal antibody (mAb).^5–9^ The public health burden of RSV is considerable, causing acute respiratory infections in individuals of all ages, contributing to over 100,000 deaths annually among children under five and significant rates of mortality among older adults.^10,11^ Thus, reliable serological evidence is needed to guide optimal vaccination strategies to reduce disease burden, particularly in children and infants.

Neutralizing antibodies (nAbs) in particular play a key role in immunity by effectively blocking viral entry into host cells and inhibiting a virus’s within-host activity, and are recognized as key serological correlates of protection for many pathogens, including RSV.^12–15^ Multiple neutralization tests, such as the plaque reduction neutralization test (PRNT), foci reduction neutralization test (FRNT), and pseudovirus neutralization test (PVNT), have been established to quantify antibody concentrations in serum.^12,16–19^ A common design for these assays involves incubating a virus with a series of diluted serum samples and measuring the experimental outcome (e.g., the number of plaques or foci of infected cells) at each dilution. As assay readouts vary across dilutions, a quantitative relationship (i.e., a dilution curve) can be inferred for each sample, serving as the basis for estimating the nAb titer, a summary statistic of the antibody concentration in a sample. Specifically, the titer is often defined as the reciprocal of the dilution that results in 50% neutralization.

Although a neutralizing antibody titer is ideally an accurate summary measurement of antibody levels in a sample from a dilution series, differences in experimental design and the absence of standard controls can limit the comparability of data across studies.^20–23^ International initiatives to standardize methods and reporting have improved inter-lab comparisons, but other sources of random and systematic error can still persist. For example, intra-laboratory variation arises from differences in reagent lots, cell passages, operators, and environmental conditions across experimental batches even if the protocol is unchanged. Moreover, even within a single batch, technical errors, such as the pipetting errors and variation in incubation conditions, can introduce uncertainty in titer estimation. Replicate measurements of the same sample are often used to average over this technical variation, but this does not adjust for systematic biases arising between batches or laboratory testing rounds.

Despite the attention paid to experimental design, less care is given to the statistical method for calculating antibody titers from dilution series data.^20^ To generate a single antibody titer estimate, a dose-response analysis is usually performed using the raw dilution series data such that the output is a single parameter or summary from a fitted model.^24^ Careful statistical analysis at this stage is arguably as important for calculating estimates such as seroprevalence or geometric mean titers (GMTs) as the experimental design, but modelling specialists are often only involved after the raw data processing stage. A number of off-the-shelf methods exist for estimating antibody titers, including the Karber formula and the four-parameter logistic (4PL) model, as well as more sophisticated, custom mathematical models.^17,24–28^ However, their performance for estimating antibody titers from raw serial dilution data has not been explored, and uncertainty in the model fits are not usually propagated into downstream analyses.

In this study, we developed a Bayesian hierarchical framework to improve antibody titer estimation from serial dilution data, generating posterior distributions for individual antibody titers while correcting for between-lab and between-batch level effects. We applied the framework to RSV FRNT data generated from a robust assay protocol with positive serum and virus controls, demonstrating how raw data generated from a standardized procedure can be paired with a sophisticated model to improve the estimation of neutralizing antibody titers. To evaluate the performance of our framework, we conducted a simulation study comparing the proposed model with standard estimation methods. We also examined how different titer estimation approaches affect population-level serological outcomes from two seroepidemiological studies in Anhua County, Hunan Province, China, including age-stratified seroprevalence, GMTs, seroconversion rates and fold-rise rates.

## Results

### Description of experimental data

This study was based on 1,777 serum samples collected between 2013 and 2021 through a cross-sectional seroepidemiological study^29^ and a mother-neonate cohort study^30^, both conducted in Anhua County, Hunan Province, China (see Supplemental Information, Section 1 for details). We performed RSV FRNTs in 28 separate batches. Each batch included three types of samples (Table S1). First, a virus control (VC) prepared at a fixed dilution of working virus stock without serum. Second, serially diluted positive controls to ensure reproducibility, including an internal positive control (PC; i.e., a pooled set of positive serum samples) and an International Standard control (IS; i.e., First WHO International Standard for Antiserum to RSV, NIBSC code: 16/284)^31^. Third, serum samples from study participants with unknown antibody concentration. The objective of the FRNT is to estimate the antibody concentration (i.e., nAb titer) in each sample. In this process, the virus control provides a measure of the background signal (i.e., the count of foci observed in the absence of serum) across different experimental conditions and batches. The positive controls, which are expected to have the same concentration in each experimental run, provide information on the effects of between-batch variation. The true concentration in each sample is therefore a latent variable of interest. However, the assay provides only noisy measurements of this latent variable. The purpose of the modelling is to obtain reliable estimates of the true antibody titers after accounting for noise and errors introduced in the measurement process (Figure. 1, see Supplemental Information, Section 2 for detailed RSV FRNT workflow).

**Figure 1.**
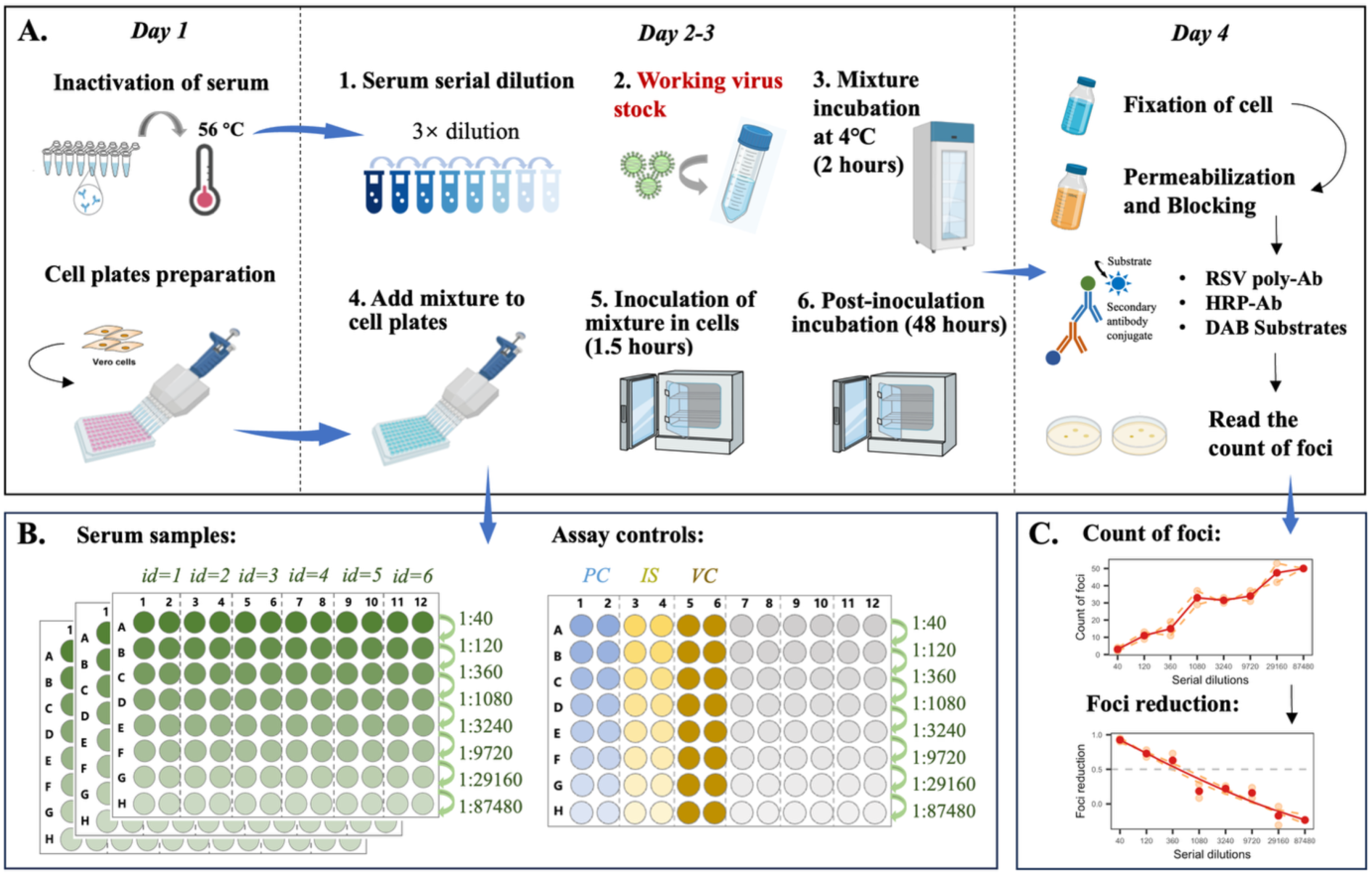
Workflow and data interpretation of the RSV FRNT assay. **(A) Overview of the RSV FRNT experimental procedure. (B) Layout of serum samples and assay controls on 96-well plates in each experimental batch.** Shading from dark to light indicates serial dilutions from high to low concentration. The VC was prepared at a fixed dilution. **(C) Example readouts showing count of foci from serially diluted serum samples, along with the fitted curve based on foci reduction.** Orange points and dashed lines represent results from two replicates; red points and solid line indicate replicate means; red curve shows the corresponding fitted curve based on the 4PL model. The intersection of the red curve with the horizontal dashed line at 50% foci reduction indicates the nAb titer estimate derived from the 4PL model.

### RSV FRNT titers show considerable variation between and within batches

The concentration of working virus stock in each well, termed the virus working dilution, directly affects foci counts in the FRNT assay. In the first 15 batches we tested virus working dilutions from 1:200 to 1:400 to identify an optimal value for countable and reproducible foci. We selected 1:330 and used this in the remaining 13 batches. Variability in VC raw measurements was observed between batches even with the same working virus stock (Figure S1A). Between-batch variability was also observed for PC and IS. In Figure S1B-C we show results from two typical batches, one with consistently higher counts of foci than the other across serial dilutions, despite both using the same working virus stock. A commonly used method for the control of batch effects is to normalize the data by calculating foci reduction at each serial dilution relative to the mean foci count of VC within the same batch. However, we still observed notable differences in both the foci reduction data and the fitted dilution curves across batches (Figure S1D-E).

We estimated the nAb titers of each PC replicate based on the Karber formula and the 4PL model. Both methods summarize the relationship between the dilution factor and the reduction in foci counts to generate a single antibody titer estimate per sample (see Supplemental Information section 3 for details; the titers were estimated based on Formula (2) and (4), respectively). In a perfect experimental and modelling setup, the measured nAb titer should be the same for all batches, as each run uses the same PC sample. However, the total variance of the 4PL titer estimates was 0.45, 30.8% of which was between-batch variance and 69.2% within-batch variance. The Karber formula resulted in even higher variability (a total variance of 0.89), with between- and within-batch variance accounting for 44.1% and 55.9%, respectively (Figure S2). Typically, nAb titers are derived from the averaged responses of multiple replicates to mitigate the impact of within-batch random errors, but the above figure refers to separate PC replicates to illustrate the potential impact of different sources of variation.

### Description of the Bayesian hierarchical model

In the above analysis we observed substantial variation in both the experimental data and the titer estimates derived from the Karber formula and 4PL model. While averaging responses across multiple replicates reduced within-batch variance, significant variability remained due to batch effects, which are often overlooked and cannot be fully addressed with standard methods such as the Karber formula and 4PL model. To provide a method that could further account for between-batch variation, we developed a Bayesian hierarchical model (BHM) based on the 4PL model. First, for each set of counts, the foci reduction was calculated relative to the modelled mean foci counts in VC for each virus working dilution. Then, we modelled the relationship between the dilution factor and reduction in foci counts using a hierarchical 4PL model, allowing for random effects both between samples and between batches. Each sample, including the PC and IS samples, has its own set of parameter values drawn from a population-level distribution. Using this hierarchical approach, we fit a 4PL curve to the dilution series data from each sample, adding both sample-level, batch-level, and optionally, experimental-level effects. Thus, the observed data are described by a 4PL curve accounting for all of these sources of variation, whereas the true, unadjusted 4PL curve for each sample is obtained by subtracting the batch- and experiment-level effects. The corrected antibody titer can then be calculated directly as a function of the unadjusted 4PL curve (Figure S3; see Supplemental Information section 4 for details. Model parameters are listed in Table S2).

### Comparison of titer estimation methods applied to experimental data

We applied the BHM to our experimental data and compared the results with those obtained from standard methods, including the Karber formula and the 4PL model. The results demonstrated that our model effectively captured the heterogeneity across batches in VC, PC, and IS (Figure 2 A-B). Comparisons of different nAb titer estimates across all tested samples showed that the Karber formula yielded the largest deviations, the 4PL model tended to produce negative estimates for samples with low antibody levels, while BHM-unadjusted posterior median estimates were symmetrically distributed around the adjusted values (Figure 2C).

**Figure 2.**
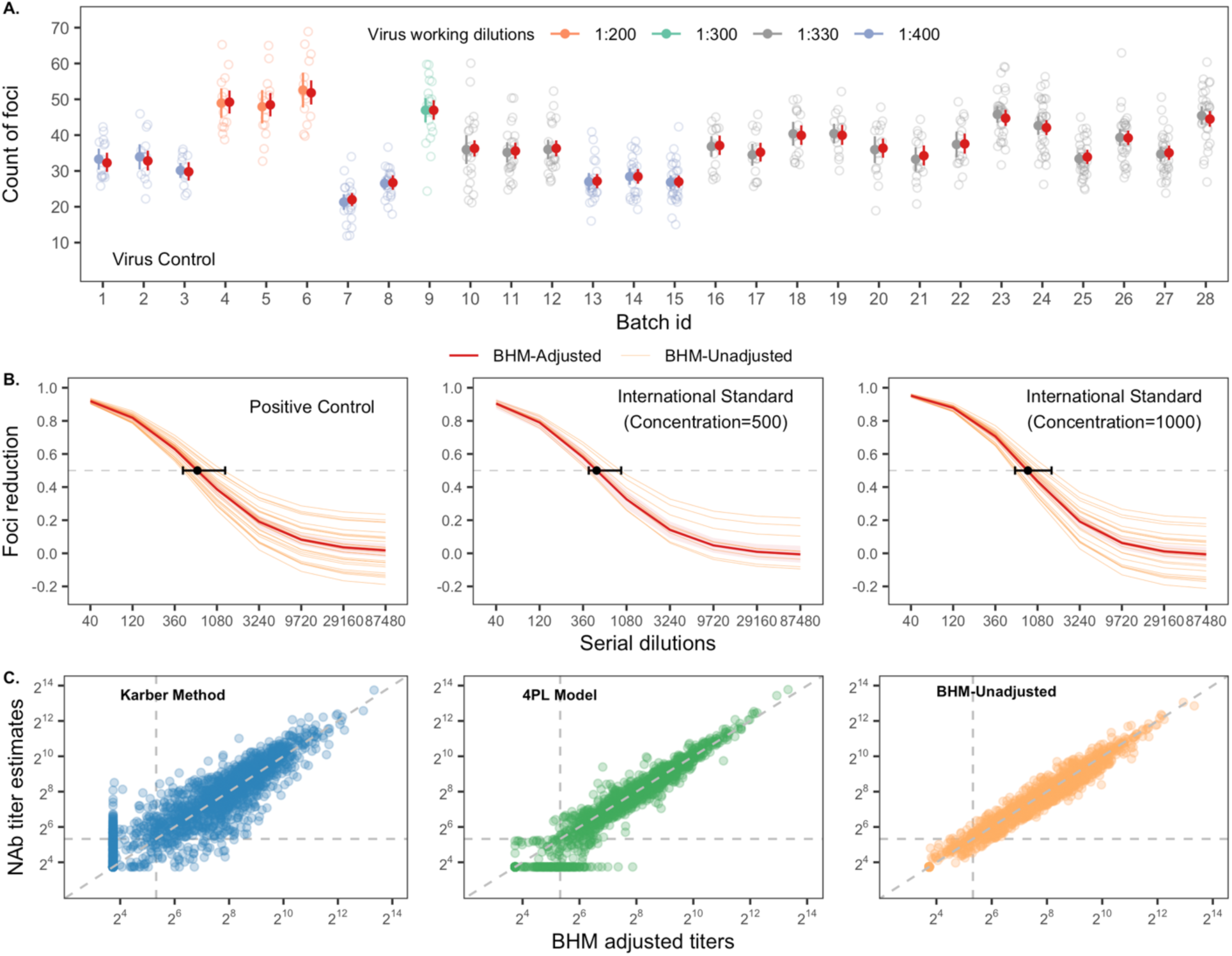
Posterior estimates from the Bayesian hierarchical modelling framework based on experimental data. **(A) Foci count in VC across batches using different working virus stocks.** Open circles represent observed counts of foci. Colored points with error bars represent the means and 95% confidence intervals of observed data. Red points with error bars represent posterior medians and 95% credible intervals. **(B) Comparison of BHM-adjusted and unadjusted curves for PC and IS across batches.** Shaded areas represent the 95% credible intervals of BHM-adjusted curves. Black points at 50% foci reduction (i.e., the grey dashed line) represent the BHM-adjusted titers for PC and IS, while error bars indicate the range of BHM-unadjusted titers across batches. **(C) Pairwise agreement between BHM-adjusted estimates and those derived from Karber formula, 4PL model, and BHM-unadjusted model.** The vertical and horizontal dashed lines indicate the seropositivity threshold.

As shown by three representative batches in Figure S4, the adjusted PC and IS titers were either lower or higher than the unadjusted ones, leading the BHM to apply corresponding downward or upward adjustments for the titer estimates of the serum samples tested within the same batches. We also found that the 4PL model sometimes failed to accurately fit curves, leading to biased nAb titer estimates. The distribution of individual nAb titer estimates were shown in Figure S5.

### Comparison of titer estimation methods applied to simulated data

We conducted a simulation study to evaluate the model performance where the true antibody titer for each sample was known (see Supplemental Information section 5 for details; simulation parameters are listed in Table S3). We generated a simulated dataset which successfully reproduced the key features of the experimental data, including the heterogeneity across working virus stocks, batches and serum samples (Figure 3). Based on the simulated VC data, we obtained reasonable estimates for the parameters of each virus working dilution (Figure 3A and Figure S6). Using these estimates, we transformed the simulated data into foci reduction to fit the BHM. We found the model closely recovered the true curves for both serum samples and assay controls (Figure 3B-C), although some parameter estimates were slightly different from their true values (Figure S7-S9). Figure 3B-C shows the true and fitted PC curves from two typical batches, along with the curves from three simulated serum samples drawn from the same batches. In these batches, we observed either higher or lower proportions of foci reduction in the PC due to batch effects. Consequently, the fitted curves, as well as the nAb titer estimates from Karber formula, 4PL model and BHM-unadjusted model, tended to be overestimated or underestimated, whereas the BHM-adjusted model consistently provided posterior median estimates closest to the true titers.

**Figure 3.**
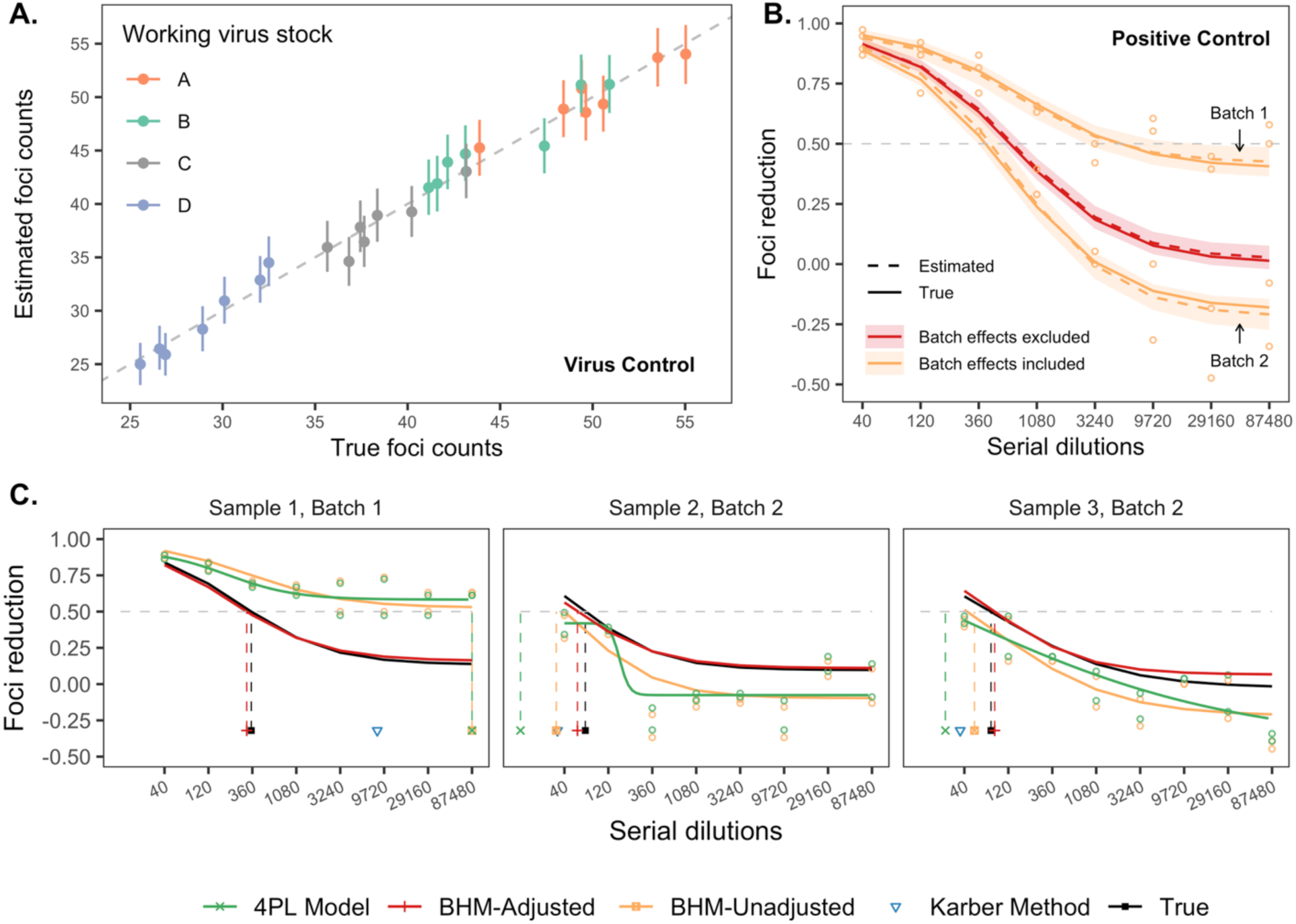
Posterior estimates from the Bayesian hierarchical modelling framework based on simulated data. **(A) Comparison of true and estimated mean foci counts in VC across all simulated batches.** Colored points with error bars represent posterior medians and 95% credible intervals. **(B) Comparison of true and fitted curves for PC in representative batches.** Shaded areas represent the 95% credible intervals. **(C) Fitted curves and nAb titer estimates for three serum samples from the same batches in (B).** Black lines indicate the simulated true curves. Green circles represent foci reduction data calculated relative to the statistical mean foci count of VC in each batch, which were used for the Karber formula and 4PL model, as defined by Supplemental Information Formula (1); Orange circles represent the same data but calculated relative to the posterior mean foci count of VC in each working virus stock, defined by Supplemental Information Formula (8). Intersections of the curves with the horizontal dashed line at 50% foci reduction mark the corresponding nAb titer estimates for each method; estimates from the Karber formula are shown as inverted triangles. Samples with fitted curves that did not achieve a 50% foci reduction were considered seronegative and had their titers imputed as the lower limit of quantification (LLOQ, i.e., 1/3 of 40). Titer estimates exceeding the final dilution point (87,480) were censored at 87,480.

We next compared the estimated nAb titers against the true values across all simulated samples (Figure 4). Among all methods evaluated, the Karber estimates exhibited the greatest uncertainty and led to increased rates of both false positives and false negatives. The 4PL and BHM-unadjusted models are more accurate than the Karber formula but are likely to produce extreme titer estimates due to large batch effects, for example, Sample 1 in Figure 3C. Additionally, the 4PL model tended to produce negative estimates in samples with low antibody levels, likely due to their inappropriate fitting patterns (e.g., Samples 2 in Figure 3C). Notably, when such low-titer samples make up a large proportion of the dataset, these errors may bias the overall titer distribution, as observed in our experimental data. The BHM-adjusted estimates, however, showed the highest accuracy. Overall, the Spearman correlations with the true titers were 0.63 (Karber estimates), 0.87 (4PL estimates), 0.84 (BHM-unadjusted estimates), and 0.96 (BHM-adjusted estimates), respectively.

**Figure 4.**
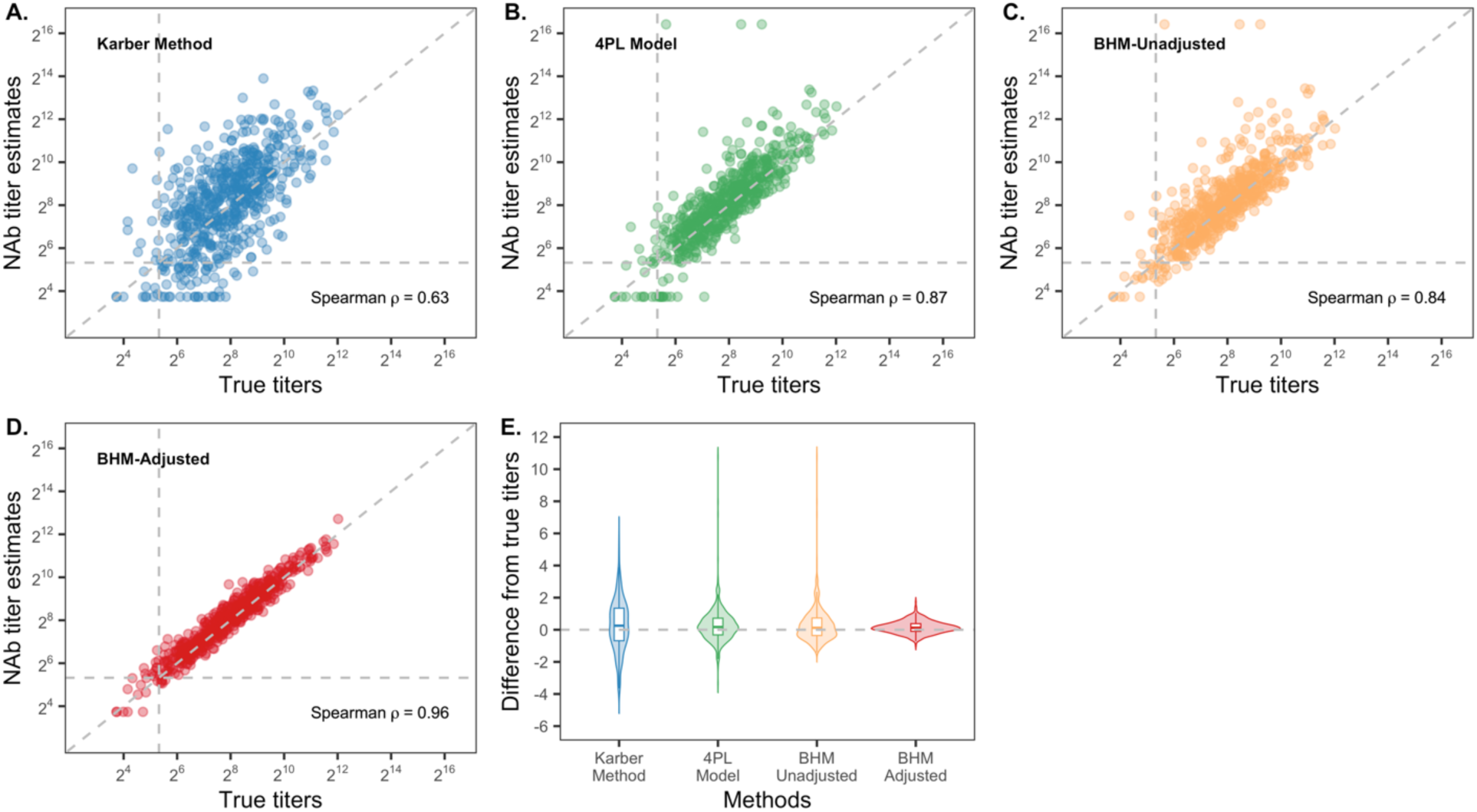
Agreement between estimated and true nAb titers across methods. **(A) Karber formula. (B) 4PL model. (C) BHM-unadjusted model. (D) BHM-adjusted model. (E) Differences between estimated and true nAb titers.** The vertical and horizontal dashed lines in (A)-(D) indicate the seropositivity threshold.

### Impact on seroepidemiology

Finally, we performed seroepidemiological analysis using different titer estimates from the experimental data. We observed notable differences across methods in the estimates of GMTs and seropositivity rates. These discrepancies were particularly evident among children aged 0-5 years, a high-risk group for severe RSV infection. For example, in infants aged 0-6 months, the GMTs estimated using the Karber formula, 4PL model, and BHM-adjusted model were 6.33 (95% confidence interval [CI]: 6.17-6.49), 5.66 (95% CI: 5.48-5.84), and 5.87 (95% CI: 5.70-6.04), respectively. The corresponding seroprevalence estimates were 66.73% (95% CI: 62.52%-70.69%), 48.23% (95% CI: 43.91%-52.57%), and 54.92% (95% CI: 50.57%-59.20%), respectively (Figure 5A-B).

**Figure 5.**
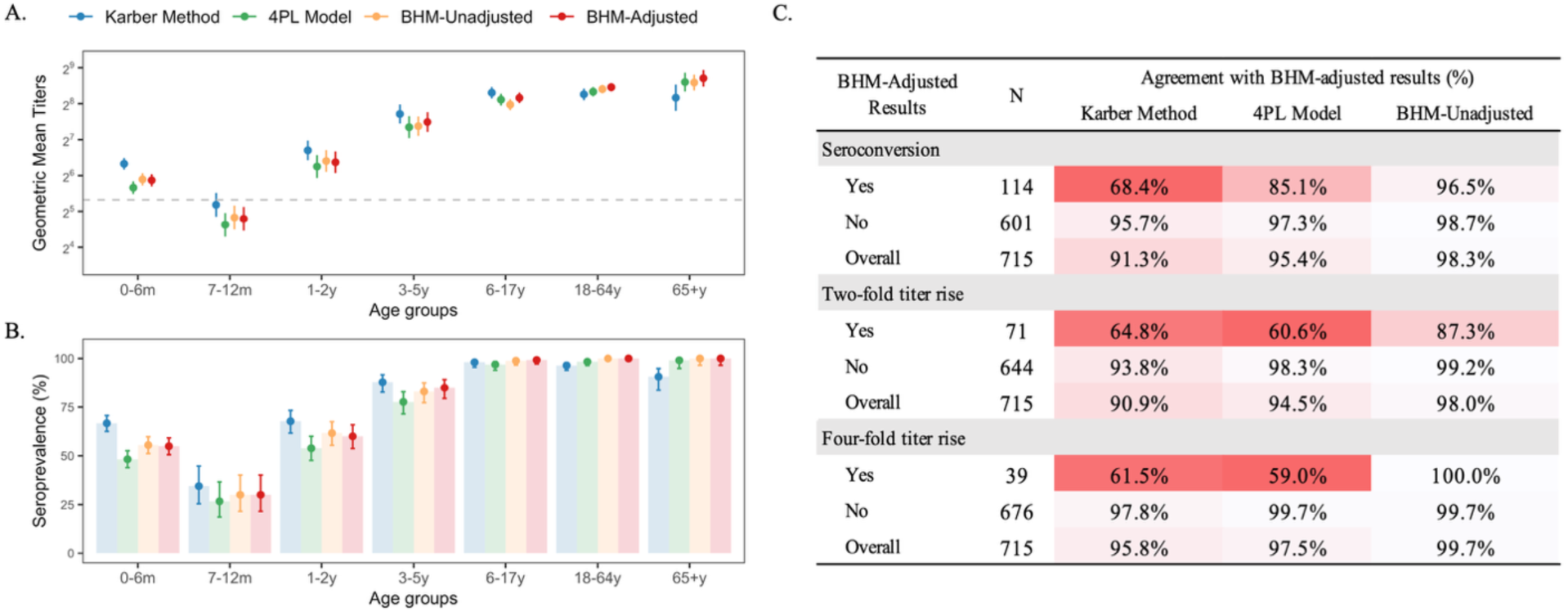
Comparison of seroepidemiological indicators based on RSV nAb titer estimates from each method. **(A-B) Age-specific GMTs and seroprevalence.** The horizontal dashed line in (A) indicates the seropositivity threshold. Error bars represent 95% confidence intervals. **(C) Agreement of the Karber formula, 4PL model, and BHM-unadjusted model with the BHM-adjusted model in identifying seroconversion and antibody fold rises in paired serum samples.** Colors from dark to light indicate increasing agreement.

Seroconversion or fold rises in nAb titers of paired serum samples are commonly used outcomes to identify new serological infections, or to assess the immunogenicity of vaccines. Based on the 715 paired serum samples from the cohort study, we used nAb titer estimates from different methods to determine whether seroconversion or a two-fold or four-fold titer rise occurred at each follow-up visit (see Supplemental Information, section 6 for details). Assuming the BHM-adjusted posterior median titer reflect the true serostatus, we found that the Karber formula showed lower sensitivity in detecting seroconversion (68.4%), two-fold titer rises (64.8%), and four-fold titer rises (61.5%). The 4PL model also showed reduced sensitivity for seroconversion (85.1%), two-fold (60.6%), and four-fold (59.0%) titer rises. Nevertheless, both methods demonstrated high specificity (>90%) across all outcomes (Figure 5C). We also observed discrepancies across methods in the population-level estimates of seroconversion rates, as well as two-fold and four-fold titer rise rates, stratified by age at each follow-up visit. Particularly, the Karber formula produced higher rates of two-fold titer rise at 6 months, 1 year, and 2 years of age. In contrast, the 4PL model yielded lower rates of both two-fold and four-fold titer rises at 1, 2, and 3 years of age (Figure S10).

## Discussion

In this study we developed a Bayesian hierarchical modelling framework based on FRNT, one of the most widely used serological assays for quantifying neutralizing antibody levels.^12^ Our primary objective was to correct for batch effects, which were found to be an influential source of variability in our experimental data but have often been overlooked in serological studies. However, the model can be adapted with minor modifications for lab- and batch-effect correction in any similar assays that produces serial dilution curves, enhancing the accuracy of serological testing. Overall, our method performed well in capturing the patterns of observational experimental data. Compared with previous approaches, the proposed model produced unbiased and more accurate estimates for both the individual-level nAb titers and the population-level seroepidemiological parameters (e.g., GMTs and seroprevalence). It could offer more reliable evidence to inform vaccine development and public health policymaking.

Both the Karber formula and 4PL model are simple and widely used methods for estimating nAb titers from raw count data in assays such as PRNT or FRNT.^17,24^ However, we found that the Karber formula can lead to high variability in individual titer estimates, potentially biasing the calculation of GMTs, seroprevalence, and other serological outcomes. We also found that the 4PL model, despite providing a flexible, model-based approach for estimating nAb titers, was suboptimal in low-titer samples, where the estimated curve often deviated substantially from the true underlying curve, leading to inaccurate titer estimates. This bias may arise because it fits each curve solely based on the data from an individual serum sample, whereas the hierarchical structure of the proposed Bayesian model provides more reasonable fits by borrowing strength across samples through partial pooling.

Inter-laboratory differences hinder comparability of titer estimates between serological studies. This arises from differences in assay protocols, reagents, and equipment.^32,33^ Although our modelling framework is capable of correcting for lab effects, controlling for the measurement errors through a standard operating procedure (SOP) remains the primary choice. Given that a standardized, universally accepted SOP for RSV FRNT assays is still lacking, in this study, we adapted the RSV FRNT assay based on an established protocol.^34^ Multiple pilot tests were performed to optimize assay procedures and ensure stability and reproducibility. The detailed methodological workflow presented in Supplemental Information section 2 provides a useful reference for future SOP development. Our incorporation of the WHO International Standard enabled comparison of our results with data from other studies. We reported an adjusted titer of 842.85 for the IS (concentration:1000 IU or 2000 IU/mL) against RSV A, suggesting that the nAb titers in this study can be converted to WHO International Units per milliliter (IU/mL) by multiplying a conversion factor of 0.421 (i.e., 842.85/2000).^35^

Intra-laboratory effects between batches, such as variation in reagent lots, operators, experimental conditions, and other random effects, are a major source of measurement error in serological assays.^36^ The WHO’s Guidelines on the Quality, Safety and Efficacy of RSV Vaccines^37^ emphasize that adequate controls should be used to standardize measurements across assay runs. These control samples also provide the basis for correcting batch effects in our Bayesian hierarchical modeling framework. In this study, random batch effects were assumed across all batches, which is reasonable given the implementation of standardized procedures: all assays were performed by the same group of trained personnel in a single laboratory, using reagents from the same manufacturer, thereby minimizing technical variation across batches. Our experimental setup had one source of systematic variation between batches in the virus working dilution, where concentrations ranging from 1:200 to 1:400 were tested to identify conditions that yielded countable and reproducible foci. We accounted for this systematic variation in the Bayesian hierarchical model (see Supplemental Information, section 4.1 for details), though this step would not be necessary if the same working virus stock were used across all FRNT batches. A similar approach could be taken in future applications to account for fixed or random effects from differences in reagents, technicians, equipment and other relevant variables across batches.

Despite the advancements of the proposed model, we recommend minimizing between-batch variation at the experimental design stage, as this reduces the need for (or further increases the performance of) complex models. The BHM-unadjusted estimates showed minimal bias, with only minor differences from the adjusted estimates (Figure 2C), likely reflecting the consistency afforded by standardized assay procedures. However, in a simulation study with amplified batch effects, BHM-unadjusted estimates deviated markedly from the true titers (Figure 4C), emphasizing the importance of batch effect correction under less controlled conditions. Furthermore, unavoidable within-batch variability, arising from sources such as pipetting errors and random measurement noise, can significantly contribute to uncertainty in the estimated titers. In this study, substantial variability was observed in repeated measurements of assay controls within the same experimental run (Figure S1-S2), emphasizing the importance of incorporating replicates to control for within-batch variability in the modelling stage.

Our study has limitations. First, the readouts of negative samples are theoretically unchanged across serial dilutions. As a result, these samples do not fit the 4PL model and were excluded from the analysis prior to fitting the BHM. Specifically, we defined ‘clear negatives’ as samples with a foci reduction of less than 30% at the starting dilution, allowing a 20% margin to account for measurement errors (see Supplemental Information, section 6 for details). Although using a mixture model with positives and negatives, and assuming bimodal parameter distributions within the BHM could potentially address this issue^38^, we chose not to implement this approach in order to reduce model complexity. Second, the four parameters of the 4PL model may be correlated. In the BHM, we did not account for potential correlations among these parameters in order to reduce model complexity. Third, we assumed that batch effects were consistent between serum samples and assay controls. However, minor discrepancies may exist, as assay controls were all placed on the last plates of each batch and may have been exposed to longer fixation and mixing times during the experimental procedure. Furthermore, due to the absence of control sera on each plate, the model cannot adjust for between-plate variability. Lastly, the BHM was applied exclusively to data generated in a single laboratory. Its robustness and generalizability require further validation using data from additional sources.

Accurate antibody titer estimates are essential for understanding the full infection burden of pathogens which are missed through routine syndromic surveillance, and for guiding effective public health interventions. In clinical trials, a significant increase in nAbs is a primary outcome for assessing vaccines or mAbs. For public health policy makers, testing for nAbs is important to determine seroprevalence, evaluate population immunity, assess real-world vaccine effectiveness and design optimal immunization strategies. Our model demonstrates improved nAb titer estimation accuracy over standard methods in both simulation studies and real-world dataset, and also provides antibody titer estimates as posterior distributions with uncertainty rather than single point estimates. This framework can be readily adapted to other serial dilution assays and has the potential to enhance the accuracy and precision of titer estimation across diverse experimental settings. Future studies should consider the importance of standardizing both experimental design and choice of mathematical model when generating serological datasets from serial dilution assays.

## Materials and Methods

Details of the materials and methods are provided in the Supplemental Information. Section 1 describes the data and sample sources. Sections 2 outlines the RSV FRNT workflow. Section 3 summarizes the standard methods used to estimate nAb titers from dilution series data, including the Karber formula and the 4PL model. Section 4 provides a detailed description of the Bayesian hierarchical modelling framework. Section 5 details the methods for statistical simulation. Section 6 contains the data processing procedures and statistical analysis for comparing different methods.

All analyses were performed in R version 4.4.0 (R Foundation for Statistical Computing, 467 Vienna, Austria, https://www.r-project.org/). The Bayesian hierarchical model was implemented using the rstan package (version 2.32.6).^39^ The 4PL model was performed using the drc package (version 3.0-1).^24,40^

## Data and code availability

Experimental data are available from the corresponding author upon reasonable request. All code used for modeling, simulation, and analysis have been deposited in the repository under https://github.com/KristyWang/BHM-RSV-nAb-titer-correction.

## Acknowledgments

We thank all participants for providing their information and serum samples for this study. We acknowledge the staff of the Anhua County Center for Disease Control and Prevention (CDC) and the township health centers at the study sites for their support with field investigations and sample collection. H.Y. is supported by the Key Program of the National Natural Science Foundation of China (82130093). J.A.H. is supported by a Wellcome Trust Early Career Award (225001/Z/22/Z). Y.W. is supported by a fellowship from the China Postdoctoral Science Foundation (2024M750556). The computations in this research were performed using the CFFF platform of Fudan University. Drawings in Figure 1A was created using BioRender.com.

## Author Contributions

H.Y. conceived and designed the study. H.Y. and J.A.H. co-supervised the project. Q.W. and M.X. established and optimized the laboratory methods. Q.W., M.X., L.Y. and J.Z. performed the laboratory experiments. L.Y. and J.Z. prepared the experimental data. Y.W., J.A.H., and C.W. developed the model. Y.W. analyzed the experimental data, conducted the simulations, and prepared the first draft of the manuscript. H.Y., J.A.H., and C.W. critically revised the manuscript for important intellectual content. All authors contributed to the review and revision of the manuscript, approved the final version as submitted, and agreed to be accountable for all aspects of the work.

## Description of Supplemental Information

Supplemental Information includes detailed materials and methods, ten figures and three tables.

