## Supplemental Information for "A modelling framework to improve antibody titer estimation from dilution series data: application to RSV Foci Reduction Neutralization Tests"

### Table of Contents

**Materials and Methods**

1. Data description
2. Foci Reduction Neutralization Test (FRNT) for RSV
3. Standard methods for estimating nAb titers from FRNT
4. Bayesian hierarchical modelling framework
5. Statistical simulation
6. Data processing procedures and statistical analysis for comparing different methods

**Figures S1-S10**

**Tables S1-S3**

**References 1-13**

#### Materials and Methods

##### 1. Data description

The serum samples analyzed in this study were derived from subsets of two investigations conducted in Anhua County, Hunan Province, China: a cross-sectional sero-epidemiological study[1] and a mother-neonate cohort study[2]. Enrollment of participants and collection of venous blood samples for the cross-sectional study were conducted between July and November 2021. The cohort study was conducted from September 2013 to November 2021. Briefly, the cohort enrolled local late-term pregnant women and their neonates at birth, with neonates subsequently followed up at 2, 4, 6, 12, and 24, 36 months, and 5-8 years of age. Peripartum venous blood samples were collected from mothers around the time of delivery, and venous blood samples were collected from child participants at baseline (cord blood at birth for neonates) and at each follow-up visit. Detailed descriptions of the two studies and the serum collection procedures have been published elsewhere[1,2].

This study was approved by the Institutional Review Board of the WHO Western Pacific Regional Office (2013.10.CHN.2.ESR), the Chinese Centre for Disease Control and Prevention (201224), the London School of Hygiene & Tropical Medicine (15698) and School of Public Health, Fudan University (IRB#2019-15-0756; #2020-11-0857; #2020-11-0857-S, #2022-02-0947 and #2022-02-0948). In the cross-sectional sero-epidemiological study, written informed consent was obtained from all participants at study enrolment. In the mother-neonate cohort study, enrolled mothers provided written informed consent for themselves and their neonates at enrollment. At each follow-up visit, informed consent forms for participants under 7 years of age were signed by their legal guardians. Participants aged between 7 and 11 years signed simplified forms, and their guardians signed the full versions. Participants older than 12 years and their guardians signed the full version of the consent form.

##### 2. Foci Reduction Neutralization Test (FRNT) for RSV

In this study, an RSV FRNT assay was established based on previously published protocols[3], with the overall workflow illustrated in Figure 1A. Briefly, Vero cells (CCL-81, American Type Culture Collection, ATCC) were seeded at a density of $2 \times 10^5$ cells per well in 96-well plates 16 hours prior to infection. Heat-inactivated serum samples were serially diluted 3-fold in minimum essential medium (MEM), ranging from 1:20 to 1:43,740. These dilutions were then mixed with an equal volume of the working solution of RSV A2 strain (ATCC), yielding final serum dilutions from 1:40 to 1:87,480 in a total volume of 240 µL per well. After incubation for 2 hours at 4 °C, the mixtures were added to the cell plates in duplicate (100 µL per well). Following a 1.5-hour incubation at 37°C in 5% $CO_2$, the inoculum was removed, and each well was overlaid with 2% carboxymethylcellulose (low viscosity, Sigma-Aldrich, St. Louis, MO) overlay medium consisting of cell culture medium supplemented with 2% fetal bovine serum (10091148, Thermo Fisher Scientific, USA) at 100 µL/well, then incubated at 37 °C for 48 hours.

The CMC overlay medium was then removed and cell monolayers were fixed with cold methanol at 4 °C for 20 minutes. Endogenous peroxidase activity was blocked for 10 minutes (P0100B; Beyotime Institute of Biotechnology, China), followed by permeabilization and blocking using 0.3% Triton X-100 and 2% goat serum in phosphate-buffered saline (PBS) for 30 minutes at room temperature. Viral foci were detected using a rabbit polyclonal antibody targeting the RSV fusion (F) glycoprotein (11049-T46, Sino Biological, China), followed by an HRP-conjugated goat anti-rabbit IgG secondary antibody (A0208, Beyotime, China). After fixation and immunostaining, the plates were scanned. The number of foci was counted using the image analyzer, EliSpot Reader (AID Diagnostika GmbH, Germany). All FRNT experiments were performed in an approved Biosafety Level 2 setting.

FRNT assays were conducted in 28 batches, with each batch comprising several 96-well plates. Each plate was used to test six serum samples, with each sample assayed in duplicate to control for random error. All assay controls were set on the last plate, including the virus control without serum (VC), an internal positive control (PC), and the WHO International Standard control (IS), which expresses the titer in International Unit (IU). The VC was tested at a fixed dilution. Both the PC and IS were subjected to the same serial dilution scheme as the test samples. The PC consisted of pooled positive sera from hospitalized RSV-infected patients and was consistently used across all batches. The IS used in this study was the first WHO International Standard for anti-RSV serum (NIBSC code: 16/284)[4]. In the first 10 batches, we used IS at a concentration of 500 IU (or 1000 IU/mL,  hereafter refer to IS500). In the last 22 batches, we used IS at a concentration of 1000 IU (or 2000 IU/mL, IS1000) including 4 batches with both IS500 and IS1000. This experimental design was intended to evaluate between-batch consistency while validating the assay's ability to detect samples with varying antibody concentrations. 500 IU also indicates a concentration of 1000 IU/mL and 1000 IU corresponds to 2000 IU/mL. Neutralization titers obtained from IS1000 were used for conversion to international units by multiplying a transition factor[5] of $2000/titer^{IS1000}$. (Figure 1B, Table S1).

To determine the nAb titer, defined as the dilution that reduces foci by 50% compared to the virus control, the number of foci counted at each serial dilution was converted into foci reduction data. Specifically, for each observation $i$ of sample $j$, the proportion of foci reduction, denoted by $y_i^{sam.j}$, is calculated as:

$$y_i^{sam.j} = 1 - \frac{C_{i|k(l)}^{sam.j}}{\bar{C}_{k(l)}^{VC}} \quad (1)$$

where $k(l)$ refers to batch $k$ with virus working dilution $l$, $C_{i|k(l)}^{sam.j}$ represents the count of foci for the $i^{th}$ observation of sample $j$ from batch $k$; $\bar{C}_{k(l)}^{VC}$ represents the mean foci count observed in the VC wells for batch $k$. (Figure 1C)

#### 3. Standard methods for estimating nAb titers from FRNT

Standard methods for estimating nAb titers from FRNT include the Karber formula and the 4PL model.[6-9] These approaches typically estimate titers using a simple formula or by independently fitting a single dose-response model to each sample's individual dilution series. Though more sophisticated, custom mathematical models have been developed that fit dilution curves in a hierarchical framework, they do not incorporate batch effects into the hierarchical structure.[10,11]

The Karber formula does not require any data smoothing or curve fitting, making it one of the simplest and most commonly used approaches for calculating FRNT-based antibody titers. The 50% endpoint titer for each sample was calculated using the following formula:

$$log_{10}(titer_{j.Karber}) = m - \Delta(\Sigma p_j - 0.5) \tag{2}$$

where $m$ is the $log_{10}$ of the highest dilution, $\Delta$ is the constant interval between serial dilutions expressed in $log_{10}$, $\Sigma p_j$ is the cumulative proportion of cells showing infection across dilutions. $\Sigma p_j$ is calculated as either $\Sigma p_j = \sum_x \bar{C}_{x|k(l)}^{sam.j} / \bar{C}_{k(l)}^{vc}$ or $\Sigma p_j = \sum_x (1 - \bar{y}_x^{sam.j})$, where $k(l)$ refers to batch $k$ with virus working dilution $l$, $x$ denotes each dilution, $\bar{C}_{x|k(l)}^{sam.j}$ and $\bar{y}_x^{sam.j}$ represent the mean count of foci and the mean foci reduction across all replicates of sample $j$ at dilution $x$, respectively.

The 4PL model is also a common way to fit experimental data generated by FRNT and thereby estimate nAb titers. Let $i$ and $j$ denote the indices of observation and sample, respectively. The form of the 4PL we use is:

$$\hat{y}_i^{sam_j} = c_j + \frac{d_j - c_j}{1 + e^{b_j\left(\log\left(x_i^{sam_j}\right) - \hat{e}_j\right)}} \tag{3}$$

where parameter $b_j$ is the slope of the 'S'-shaped curve; parameters $c_j$ and $d_j$ are lower and upper asymptotes of the curve, respectively; $\hat{e}_j$ is the log-transformed midrange dilution. $\hat{y}_i^{sam_j}$ is the expected proportion of foci reduction at dilution $x_i^{sam\_j}$. The nAb titer of sample $j$ is then estimated based on the dilution at 50% foci reduction:

$$\log(titer_{j.4PL}) = \frac{\log\left(\frac{d_j - c_j}{0.5 - c_j}\right) - 1}{b_j} + \hat{e}_j \tag{4}$$

We fitted the 4PL Model for each sample using the drm function from the drc package (version 3.0-1) in R (version 4.4.0).[7,12]

#### 4. Bayesian hierarchical modelling framework

In this study, we developed a Bayesian hierarchical modelling framework to correct for experimental and batch effects in RSV FRNT assays and to estimate adjusted nAb titers, based on the above 4PL model.

##### 4.1 Estimating the effects of each virus working dilution

Let $i$ denote the observation index, $k$ denote the batch, and $l$ denote the virus working dilution. We modelled the count of foci observed in each VC well, denoted by $C_{i|k(l)}^{VC}$, as Poisson distributed:

$$C_{i|k(l)}^{VC} \sim Poisson\left(\mu_{k(l)}^{VC}\right) \tag{5}$$

where $\mu_{k(l)}^{VC}$ indicates the mean count of foci for VC wells from batch $k$ at virus working dilution $l$. We modelled heterogeneity in $\mu_{k(l)}^{VC}$ as arising from two sources: a fixed effect for the virus working dilution, denoted by $\delta_l$, and a random effect for the batch, denoted by $\varsigma_k$. We first built a Bayesian hierarchical model to provide robust estimates of the mean foci count for each working virus stock ($\delta_l$). The model is specified as:

$$\mu_{k(l)}^{VC} = \delta_l + \varsigma_k \tag{6}$$

$$\varsigma_k \sim N\left(0, \sigma_\varsigma\right) \tag{7}$$

Similar to the definition in Formula (1), where the foci reduction was calculated as the foci count at each serial dilution relative to the mean foci count of VC in the corresponding batch, in our model, it was calculated relative to mean foci count of VC for the corresponding virus working dilution (i.e., the estimated parameter $\delta_l$), given as follows:

$$y_{i|k}^{T} = 1 - \frac{C_{i|k(l)}^{T}}{\delta_l} \tag{8}$$

where $T$ indicates the type of sample, i.e, $T = PC$, $IS500$, $IS1000$ or $sam.j$.

##### 4.2 Estimating the random batch effects

Next, we estimated the batch-specific effects based on the foci reduction data from the PC and IS. We modelled the foci reduction data as following a normal distribution. The model is specified as:

$$y_{i|k}^{T} \sim N\left(\mu_{i|k}^{T}, \sigma_y\right) \tag{9}$$

$$\mu_{i|k}^{T} = f(x_{i|k}^{T}, \theta_k^{T}) \tag{10}$$

$$f(x_{i|k}^{T}, \theta_k^{T}) = c_k^{T} + \frac{d_k^{T} - c_k^{T}}{1 + e^{b_k^{T}\left(\log\left(x_{i|k}^{T}\right) - \hat{e}_k^{T}\right)}} \tag{11}$$

Where $T$ indicates the type of control samples, i.e, $T = PC$, $IS500$ or $IS1000$. $x_{i|k}^{T}$ denotes the dilution corresponding to each observation, $f(\cdot)$ denotes the 4PL function and $\theta_k^{T} = [b_k^{T}, c_k^{T}, d_k^{T}, \hat{e}_k^{T}]'$ represents the set of unadjusted parameters for batch $k$ and ' denotes transpose. We modelled batch effects as having an additive effect on $\theta_k^{T}$:

$$\theta_k^{T} = \theta^{T} + \gamma_k \tag{12}$$

where $\theta^{T} = [b^{T}, c^{T}, d^{T}, \hat{e}^{T}]'$ are the mean parameters for the control sera over many batches, and $\gamma_k = [\gamma_{b,k}, \gamma_{c,k}, \gamma_{d,k}, \gamma_{\hat{e},k}]'$ denotes the random effects specific to batch $k$. We modelled the batch effect for each parameter as independently normally distributed:

$$\gamma_{b,k} \sim N\left(0, \sigma_{\gamma_b}\right) \tag{13}$$

$$\gamma_{c,k} \sim N\left(0, \sigma_{\gamma_c}\right) \tag{14}$$

$$\gamma_{d,k} \sim N\left(0, \sigma_{\gamma_d}\right) \tag{15}$$

$$\gamma_{\hat{e},k} \sim N\left(0, \sigma_{\gamma_{\hat{e}}}\right) \tag{16}$$

##### 4.3 Estimating the adjusted titer for each serum sample

Finally, the model was applied to the experimental data from the serum samples. We modelled the observed foci reduction data $y_{i|k}^{sam.j}$ as:

$$y_{i|k}^{sam.j} \sim N\left(\mu_{i|k}^{sam.j}, \sigma_y\right) \tag{17}$$

$$\mu_{i|k}^{sam.j} = f(x_{i|k}^{sam.j}, \theta_{j|k}^{*}) \tag{18}$$

$$f(x_{i|k}^{sam.j}, \theta_{j|k}^{*}) = c_{j|k}^{*} + \frac{d_{j|k}^{*} - c_{j|k}^{*}}{1 + e^{b_{j|k}^{*}\left(\log\left(x_{i|k}^{sam.j}\right) - \hat{e}_{j|k}^{*}\right)}} \tag{19}$$

where $\theta_{j|k}^{*} = [b_{j|k}^{*}, c_{j|k}^{*}, d_{j|k}^{*}, \hat{e}_{j|k}^{*}]'$ represents the set of unadjusted parameters for sample $j$ from batch $k$. We modelled each parameter as independently normally distributed:

$$b_{j|k}^{*} \sim N(b^{pop}, \sigma_b) \tag{20}$$

$$c_{j|k}^{*} \sim N(c^{pop}, \sigma_c) \quad (21)$$

$$d_{j|k}^{*} \sim N(d^{pop}, \sigma_d) \quad (22)$$

$$\hat{e}_{j|k}^{*} \sim N(\hat{e}^{pop}, \sigma_{\hat{e}}) \quad (23)$$

where $\theta^{pop} = [b^{pop}, c^{pop}, d^{pop}, \hat{e}^{pop}]'$ denotes population-level mean parameters across all serum samples, and $\sigma_b$, $\sigma_c$, $\sigma_d$, $\sigma_{\hat{e}}$ are the corresponding standard deviations. To account for both sample-level and batch-level heterogeneity embedded in $\theta_{j|k}^{*}$, we derived the adjusted sample-specific parameters $\theta_j = [b_j, c_j, d_j, \hat{e}_j]'$ by removing the estimated batch effects $\gamma_k$ (estimated from PC and IS) from $\theta_{j|k}^{*}$:

$$\theta_j = \theta_{j|k}^{*} - \gamma_k \quad (24)$$

The adjusted nAb titers were then calculated based on the batch-corrected parameters $\theta_j$ as:

$$log(titer_{j.BHM.adj}) = \frac{\log\left(\frac{d_j - c_j}{0.5 - c_j}\right) - 1}{b_j} + \hat{e}_j \quad (25)$$

For comparison, we also calculated the unadjusted titers based on the unadjusted parameters $\theta_{j|k}^{*}$:

$$log(titer_{j.BHM.unadj}) = \frac{\log\left(\frac{d_{j|k}^{*} - c_{j|k}^{*}}{0.5 - c_{j|k}^{*}}\right) - 1}{b_{j|k}^{*}} + \hat{e}_{j|k}^{*} \quad (26)$$

We performed Bayesian inference using the RStan package (version 2.32.6)[13] running four parallel Markov chains across four computational cores. Each chain was run for 6,000 iterations, including 3,000 warm-up iterations for algorithm adaptation and 3,000 post-warm-up iterations retained for posterior sampling. We set the target acceptance probability to 0.95 to improve the stability of the Hamiltonian Monte Carlo (HMC) algorithm and reduce the likelihood of divergent transitions. We employed non-centered parameterizations for all population-level, batch-level, and sample-level parameters to improve sampling efficiency. Detailed notations and prior distributions are provided in Table S2.

#### 5. Statistical simulation

We conducted a simulation study to recover the structure of RSV FRNT experimental data and assess the model performance. To match the real data, we simulated four levels of virus working dilution, with the mean foci count in the virus control to be $\delta = [\delta_1, \delta_2, \delta_3, \delta_4]'$. For each level of virus working dilution, seven experimental batches were simulated. In each batch, 20 serum samples, an internal positive control, an International Standard control (at different concentrations, i.e., IS500 and IS1000), and a virus control were included. Each serum sample, along with the PC and IS, was tested in duplicate across 3-fold serial dilutions ranging from 1:40 to 1:87480. For the VC, 22 replicates were simulated per batch.

The 4PL parameters for PC, IS500 and IS1000 (denoted by $\theta^{PC}, \theta^{IS500}$and $\theta^{IS1000}$, respectively), as well as the population-level mean parameters for serum samples (denoted by $\theta^{pop}$) were set in the simulation to match the posterior median estimates from the real data. Batch-level random effects on each 4PL parameter (denoted by $\gamma_b, \gamma_c, \gamma_d$ $and$ $\gamma_{\hat{e}}$) and the effect on VC foci counts ($\varsigma$) were collected into a vector (denoted by $\Gamma$) and drawn from a multivariate normal distribution. Similarly, sample-level random effects (denoted by $\lambda_b, \lambda_c, \lambda_d$ $and$ $\lambda_{\hat{e}}$, collected into a vector denoted by $\Lambda$) were also drawn from a multivariate normal distribution. All parameter settings for simulation are listed in Table S3.

The observed foci count for each well was drawn from a Poisson distribution:

$$C^T_{i|k(l)} \sim Poisson\left(\mu^T_{i|k(l)}\right) \tag{27}$$

where $T$ indicates the type of sample.

For each VC well (i.e., $T = VC$),

$$\mu^T_{i|k(l)} = \delta_l + \varsigma_k \tag{28}$$

For each PC or IS well (i.e., $T = PC$, $IS500$ or $IS1000$) at a given dilution $x$,

$$\mu^T_{i|k(l)} = \delta_l\left(1 - f(x^T_{i|k(l)}, (\theta^T + \gamma_k))\right) \tag{29}$$

where $f(\cdot)$ denotes the 4PL function, and $\gamma_k = [\gamma_{b,k}, \gamma_{c,k}, \gamma_{d,k}, \gamma_{\hat{e},k}]'$ represents the batch effect.

For each well of serum sample $j$ (i.e., $T = sam.j$) at a given dilution $x$:

$$\mu^T_{i|k(l)} = \delta_l\left(1 - f\left(x^T_{i|k(l)} | \theta^{pop} + \lambda_j + \gamma_k\right)\right) \tag{30}$$

where $\lambda_j = [\lambda_{b,j}, \lambda_{c,j}, \lambda_{d,j}, \lambda_{\hat{e},j}]'$ represents the sample-level random effect.

The 4PL parameters and the true titer for each simulated serum sample $j$ were defined as:

$$\theta_j = \left[b_j, c_j, d_j, \hat{e}_j\right] = \theta^{pop} + \lambda_j \tag{31}$$

$$log\left(titer_{j.true}\right) = \frac{\log\left(\frac{d_j - c_j}{0.5 - c_j}\right) - 1}{b_j} + \hat{e}_j \tag{32}$$

We applied the standard methods, including the Karber formula and the 4PL model, as well as the BHM to the simulated data. For the BHM, the posterior median was used as the titer estimate for each sample. Model performance was assessed by comparing the nAb titer estimates from each method with the true simulated titers. The consistency of the population-level, batch-level and sample-level parameter estimates from the BHM with the true values was also evaluated.

#### 6. Data processing procedures and statistical analysis for comparing different methods

In this study, seropositivity threshold was defined as a titer of 40, corresponding to the starting dilution. Seroprevalence was defined as the proportion of individuals with nAb titers at or above the predefined seropositivity threshold. Seroconversion was defined as a change from seronegative to seropositive status in paired serum samples. Two-fold and four-fold titer rises were defined as at least a twofold or fourfold increase in nAb titers between paired samples. The lower limit of quantification (LLOQ) was set at one-third of the starting dilution (i.e., 1/3 of 40). “Clear negative” samples were defined as those exhibiting less than 30% foci reduction at the starting dilution, allowing a 20% margin to account for potential measurement error due to intra- or inter-batch variation. This is reasonable as the foci reduction data decreases monotonically with dilution, indicating that the true curves for such samples are highly unlikely to reach the 50% foci reduction threshold.

For the Karber formula, titer estimates falling below LLOQ were censored at the LLOQ. For methods based on the 4PL model, including both the standard 4PL and the BHM, "clear negative" samples were excluded from curve fitting and assigned a titer equal to the LLOQ. Likewise, samples with fitted curves that did not achieve a 50% foci reduction were considered seronegative and had their titers imputed as the LLOQ. For all methods, titer estimates exceeding the final dilution point (87,480) were censored at 87,480.

We used a generalized linear model (GLM) to characterize the variation in RSV FRNT nAb titers estimated by the Karber formula and the 4PL model. We calculated Spearman correlation coefficients to assess the agreement between the estimated and true simulated nAb titers. To examine how different titer estimation approaches affect population-level serological outcomes, we calculated age-specific geometric mean titers (GMTs) and seroprevalence, as well as the corresponding 95% confidence interval (95% CI), based on the individual titer estimates from each method. Although BHM generates a posterior distribution of the nAb titer for each sample, we used only the posterior median as a single-point individual titer estimate for calculating the population-level indicators. Using paired serum samples from two consecutive follow-up visits in the cohort study, we further evaluated the consistency among different titer estimation methods in identifying serologic evidence of new infections. Specifically, we assessed their agreement in detecting seroconversion, two-fold titer rise and four-fold titer rise. Assuming the BHM-adjusted estimates reflect the true serostatus, sensitivity was defined as the proportion of individuals classified as having seroconversion, two-fold titer rise, or four-fold titer rise by a given method among those identified as such by the BHM-adjusted estimates. Specificity was defined as the proportion of individuals classified as not having seroconversion, two-fold titer rise, or four-fold titer rise by the method among those similarly classified by the BHM-adjusted estimates. Overall agreement was defined as the proportion of individuals whose classification (presence or absence of serologic change) matched that of the BHM-adjusted estimates.

#### Supplemental figures

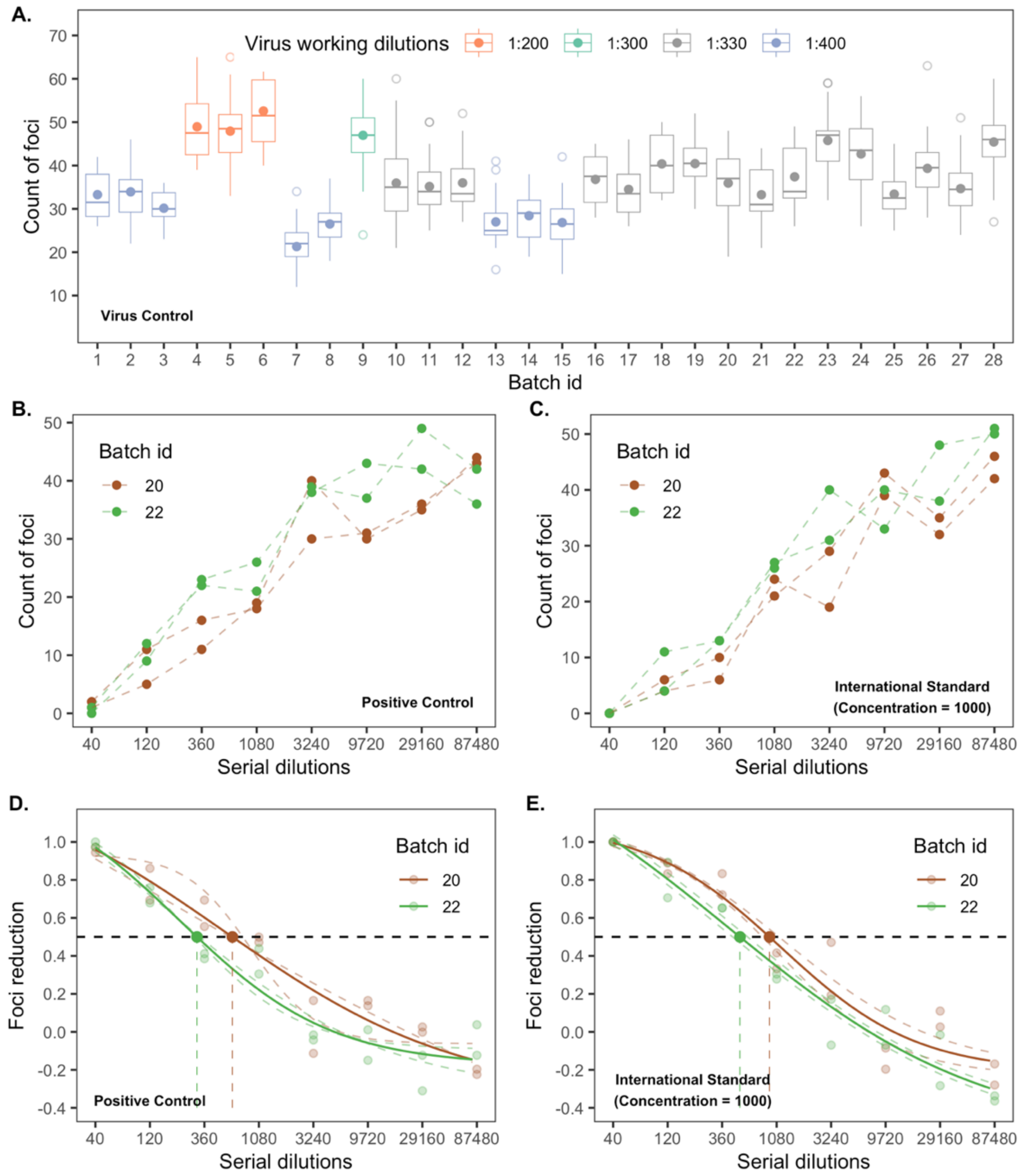


**Figure S1. Uncertainty in RSV FRNT readouts and 4PL model fittings. (A) Foci count of VC across all batches.** Solid points represent mean foci count for each batch; open circles indicate outliers. **(B) Foci count across serial dilutions of PC from two representative experimental batches.** Dashed lines indicate two replicates within each batch. **(C) Same as (B), but for IS1000. (D) Fitted 4PL curves for PC from the same batches as (C).** Foci reduction was calculated relative to the mean foci counts of VC in each batch, as defined by Formula (1). Dashed curves represent fits to individual replicates; solid curves indicate fits to replicate means. Points at 50% foci reduction (i.e., the black dashed line) denote the estimated nAb titers. **(E) Same as (D), but for IS1000.**

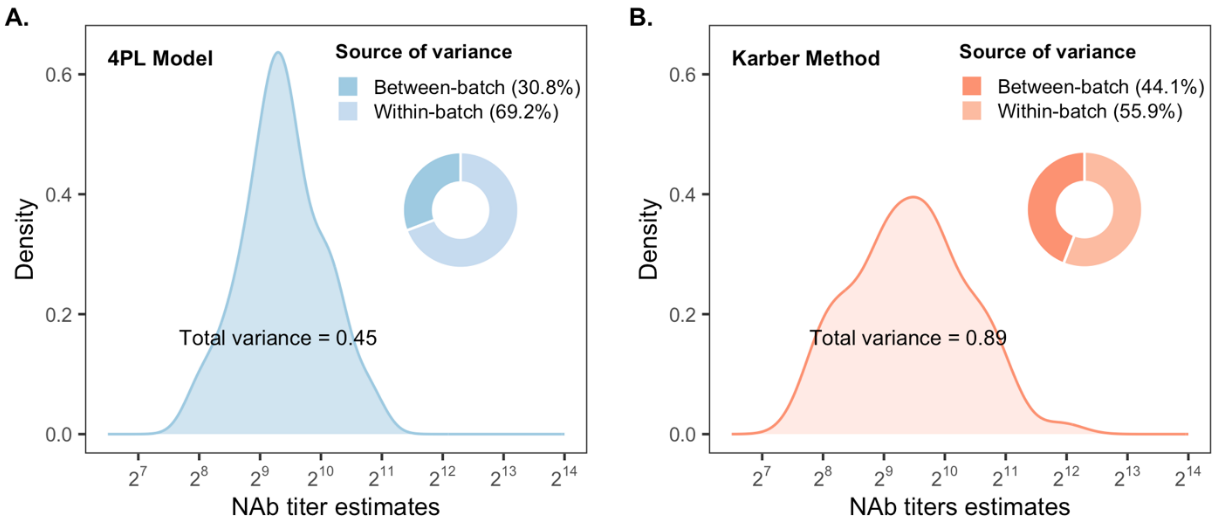


**Figure S2. Uncertainty in RSV nAb titers for the PC estimated by (A) 4PL model and (B) Karber formula.** The density plots illustrate the distribution of titer estimates, while the donut charts show the relative contributions of between- and within-batch sources of variance.

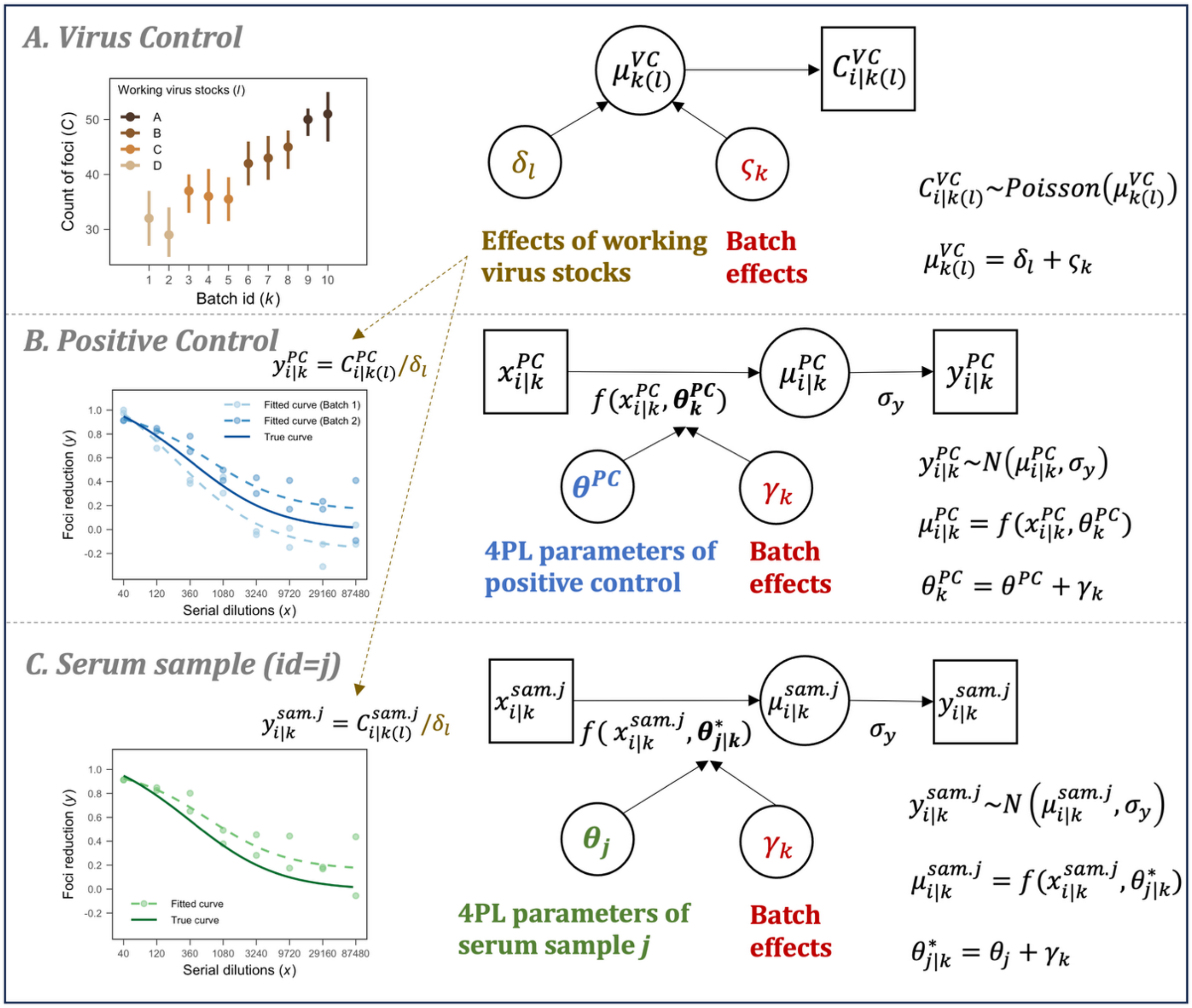


**Figure S3. Bayesian hierarchical modeling framework for correcting batch effects in RSV FRNT measurements.** Panels from top to bottom show representative experimental data (left) and corresponding schematic model diagrams (right) used to fit VC, PC, and serum sample data, respectively. The model diagram used for fitting IS is the same as that used for the PC. Squares represent observed variables, circles indicate model parameters, and arrows denote conditional dependencies. Model notation is defined in Materials and Methods, Section 4 and Table S2.

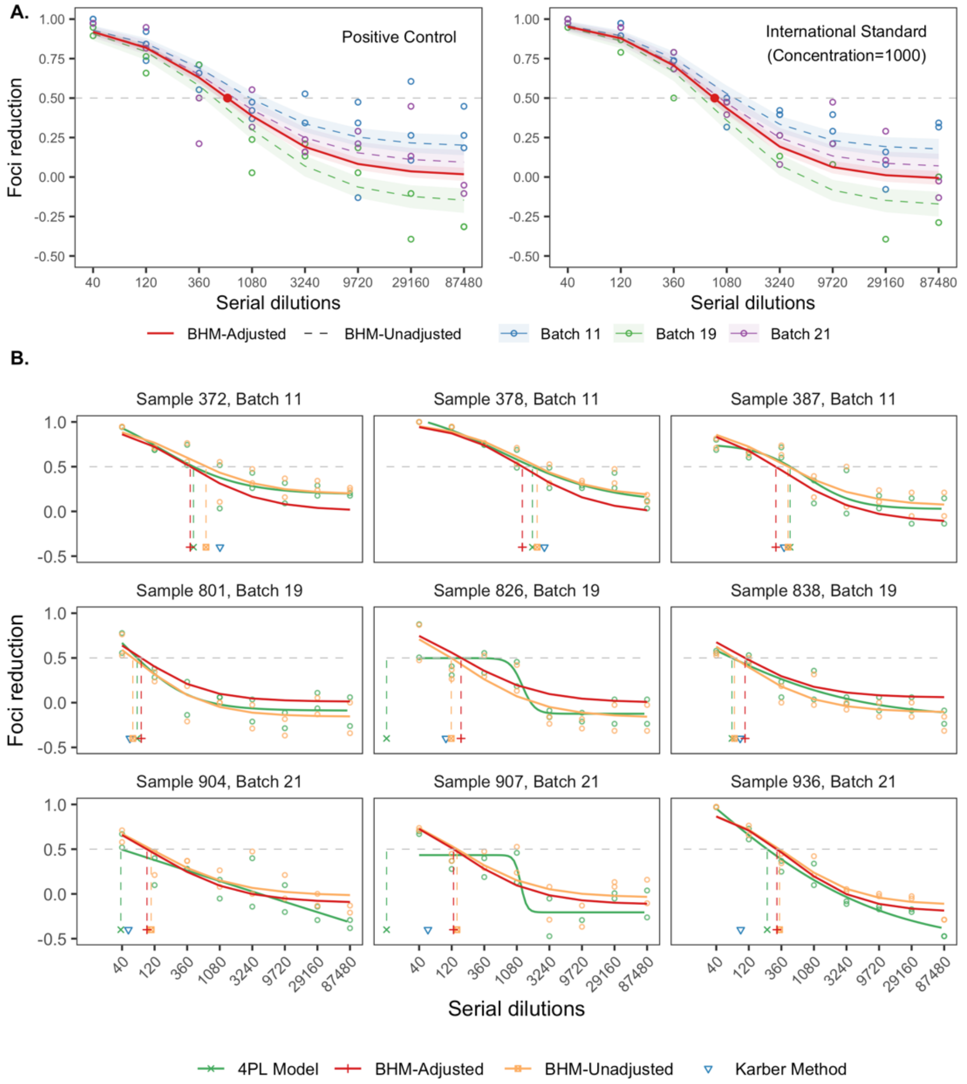


**Figure S4. Fitted curves and nAb titer estimates using different methods for PC and IS (A), and for representative samples (B) from three typical batches.** Colored circles in (A) and orange circles in (B) represent foci reduction data calculated relative to the posterior mean foci count of VC in each working virus stock, as defined by Formula (8); Green circles in (B) represent the same data but calculated relative to the statistical mean foci count of VC in each batch, which were used for the Karber formula and 4PL model, as defined by Formula (1). Shaded areas represent the 95% credible intervals. Intersections of the curves with the horizontal dashed line at 50% foci reduction mark the corresponding nAb titer estimates for each method; estimates from the Karber formula are shown as inverted triangles. Samples with fitted curves that did not achieve a 50% foci reduction were considered seronegative and had their titers imputed as the lower limit of quantification (LLOQ, i.e., 1/3 of 40). Titer estimates exceeding the final dilution point (87,480) were censored at 87,480. It should be noted that the 4PL model failed to accurately fit curves for Sample 826 and 907.

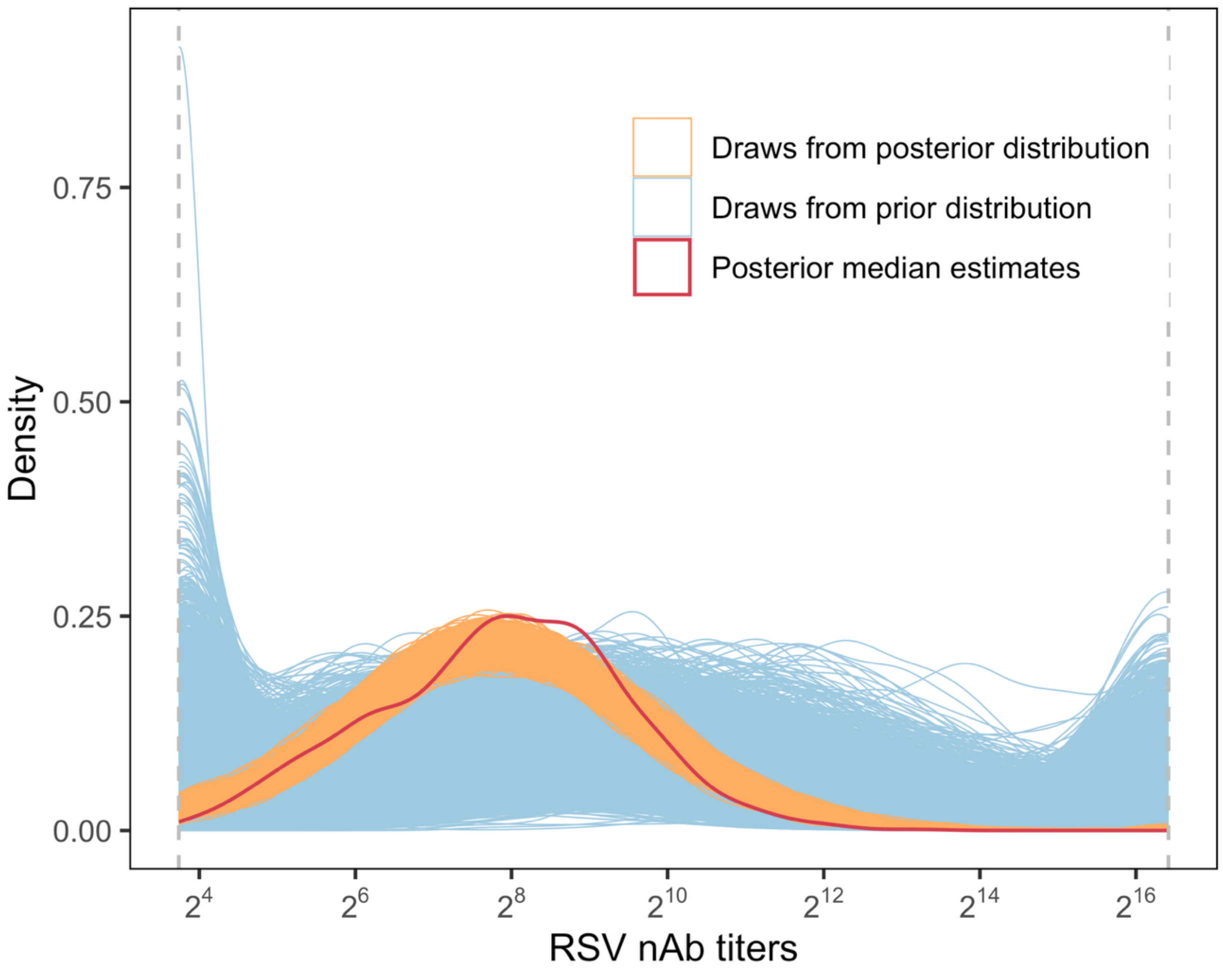


**Figure S5. Model-predicted distribution of individual nAb titers at the population level.** The red line represents the distribution of posterior median titer estimates across all tested samples (excluding “clear negative” samples, as defined in Materials and Methods, Section 6). The vertical dashed lines indicate the lower and upper limits of titer quantification (see Materials and Methods, Section 6 for details). Each remaining line represents a titer distribution defined by parameter estimates drawn from either the prior (blue) or posterior (orange) distributions. Specifically, for each draw, we simulated 1,400 new samples using the estimates of the population-level parameters based on Formulas (20)-(23). Each new sample was randomly assigned to one of 28 batches (50 individuals per batch). The adjusted sample-specific parameters were obtained for each sample based on Formula (24), and an adjusted nAb titer can then be calculated using Formula (25). This yielded one realization of the distribution of individual nAb titers per posterior draw. We show 3,000 draws each from the prior and posterior distributions.

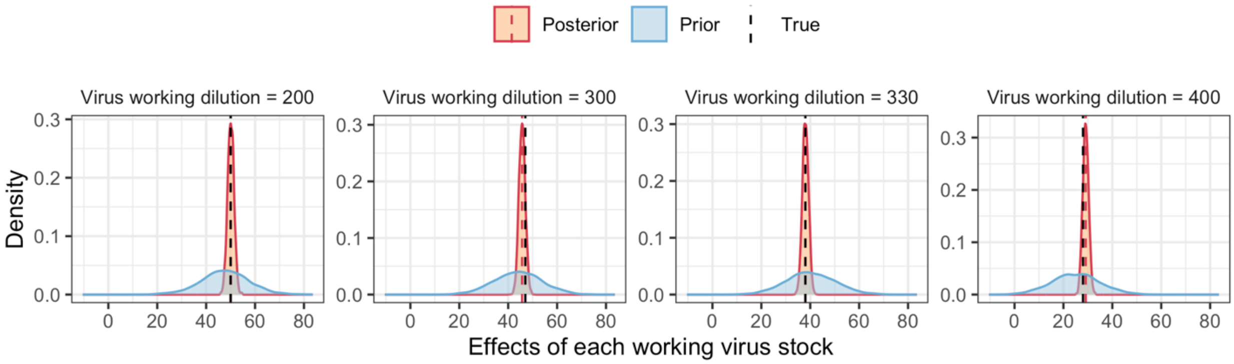


**Figure S6. Prior and posterior distributions of virus working dilution effects (denoted by $\boldsymbol{\delta}$) based on the simulated data.** Black dashed lines indicate true parameter values used in the simulation; red dashed lines show BHM posterior medians.

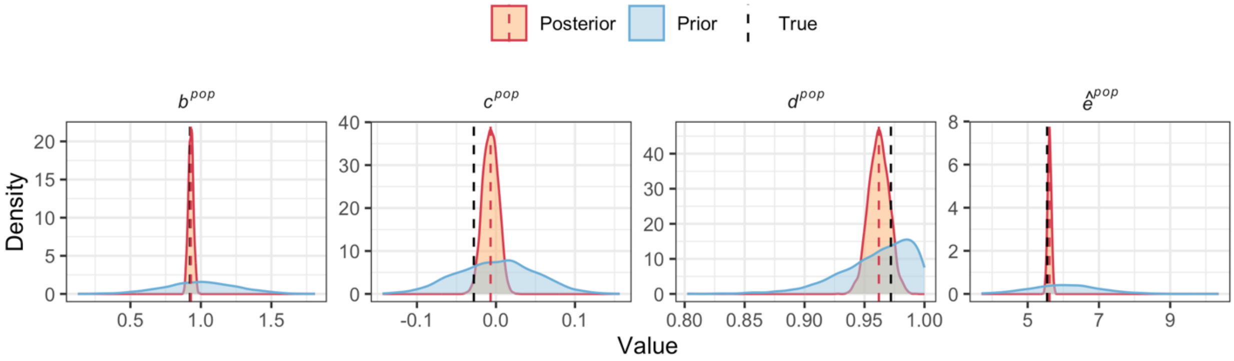


**Figure S7. Prior and posterior distributions of population-level 4PL parameters (denoted by $\boldsymbol{\theta^{pop}}$) based on the simulated data.** Black dashed lines indicate true parameter values used in the simulation; red dashed lines show BHM posterior medians.

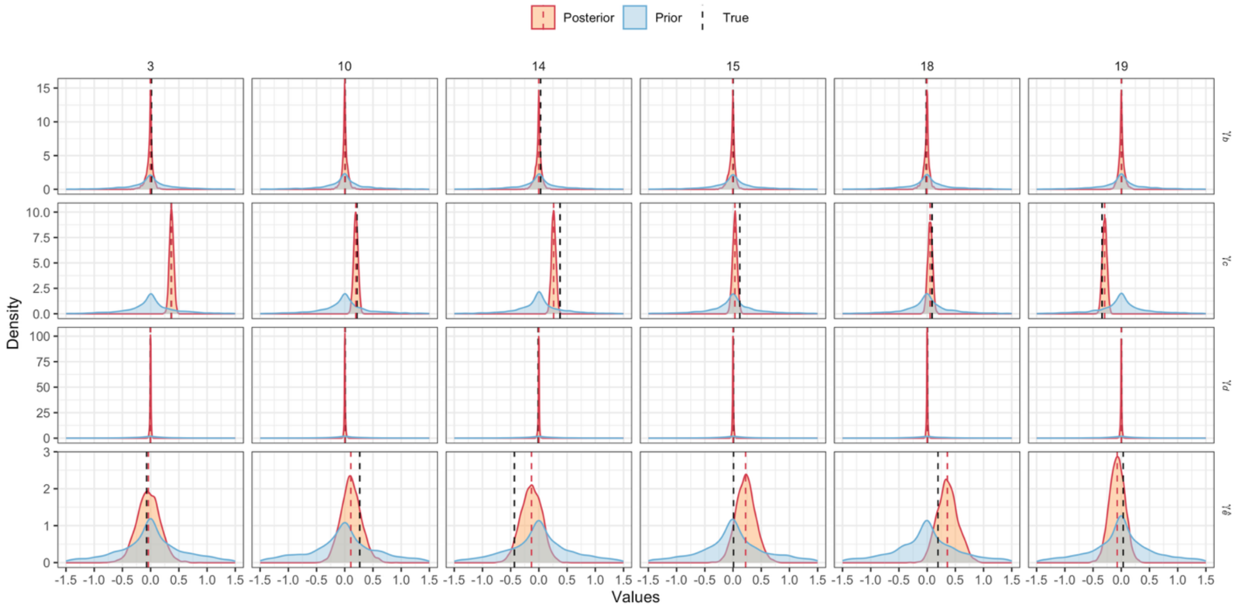


**Figure S8. Prior and posterior distributions of batch-level random effects (denoted by $\gamma_k$) for six randomly selected simulated batches.** Black dashed lines indicate true parameter values used in the simulation; red dashed lines show BHM posterior medians.

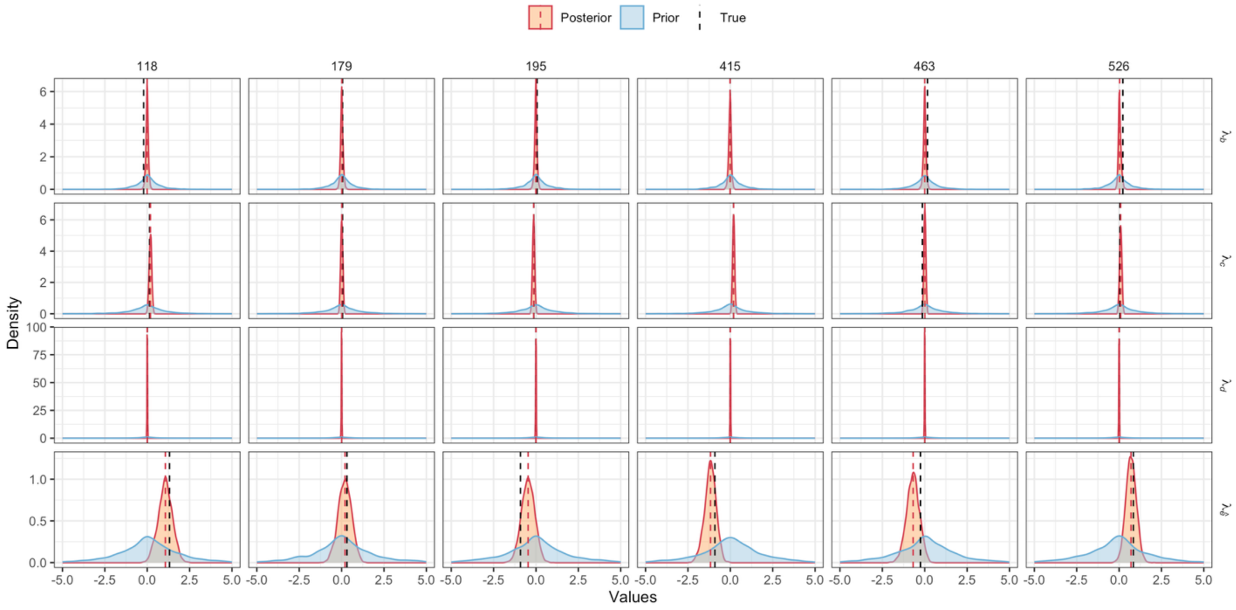


**Figure S9. Prior and posterior distributions of sample-level random effects (denoted by $\lambda_k$) for six randomly selected simulated samples.** Black dashed lines indicate true parameter values used in the simulation; red dashed lines show BHM posterior medians.

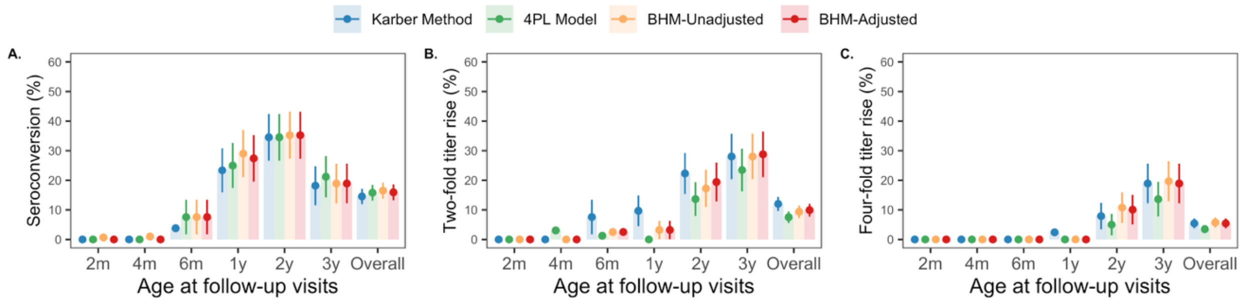


**Figure S10. Comparison of different methods in estimating seroconversion rates (A), two-fold titer rise rates (B) and four-fold titer rise rates (C) in paired serum samples.** Error bars represent 95% confidence intervals.

#### Supplemental tables

**Table S1. Summary of RSV FRNT experiments in assay controls.**

| Characteristics | Assay Controls | | | |
|---|---|---|---|---|
| | VC | PC | IS500 | IS1000 |
| Number of batches | 28 | 28 | 10 | 22 |
| Number of replicates per batch | 14-32 | 2-4 | 2-3 | 2-4 |
| Total number of replicates | 603 | 67 | 24 | 65 |

**Table S2. Notations for data variables and prior distributions for model parameters.**

| Notation | Description | Prior distribution[+] |
|---|---|---|
| **Indexing variables** | | |
| $i$ | Index for observation | - |
| $j$ | Index for serum sample | - |
| $k$ | Index for batch | - |
| $l$ | Index for working virus stock | - |
| **Data variables** | | |
| $x$ | Dilution | - |
| $C$ | Count of foci | - |
| $y$ | Proportion of foci reduction | - |
| **Model parameter** | | |
| $\delta_l$ | Mean count of foci in VC for working virus stock $l$[†] | $\delta_1 \sim N(48, 10)$<br>$\delta_2 \sim N(44, 10)$<br>$\delta_3 \sim N(40, 10)$<br>$\delta_4 \sim N(25, 10)$ |
| $\varsigma_k$ | Deviation in the mean foci count of VC of batch $k$ from $\delta_l$ | $\varsigma_k \sim N(0, \sigma_\varsigma)$<br>$\sigma_\varsigma \sim N(0, 1)[0, ]$ |
| $\theta^{pop}$ | Population-level mean parameters across all serum samples[‡] | $b^{pop} \sim N(1, 0.25)[0, 2]$<br>$c^{pop} \sim N(0, 0.05)[-1, 1]$<br>$d^{pop} \sim N(1, 0.05)[0, 1]$<br>$\hat{e}^{pop} \sim N(6, 1)[3.7, 11.4]$ |
| $\theta^{PC}$ | 4PL parameters for PC | Same as $\theta^{pop}$ |
| $\theta^{IS500}$ | 4PL parameters for IS500 | Same as $\theta^{pop}$ |
| $\theta^{IS1000}$ | 4PL parameters for IS1000 | Same as $\theta^{pop}$ |
| $\gamma_k$ | Batch-specific random effects. | $\gamma_{b,k} \sim N(0, \sigma_{\gamma_b})$; $\sigma_{\gamma_b} \sim N(0, 0.5)[0, ]$<br>$\gamma_{c,k} \sim N(0, \sigma_{\gamma_c})$; $\sigma_{\gamma_c} \sim N(0, 0.5)\ [0, ]$<br>$\gamma_{d,k} \sim N(0, \sigma_{\gamma_d})$; $\sigma_{\gamma_d} \sim N(0, 0.5)[0, ]$<br>$\gamma_{\hat{e},k} \sim N(0, \sigma_{\gamma_{\hat{e}}})$; $\sigma_{\gamma_{\hat{e}}} \sim N(0, 1.0)[0, ]$ |
| $\theta^*_{j\|k}$ | Sample-specific 4PL parameters without adjusting for batch effects | $b^*_{j\|k} \sim N(b^{pop}, \sigma_b)$; $\sigma_b \sim N(0,0.5)[0, ]$ |

| Notation | Description | Prior distribution[+] |
|---|---|---|
| | | $c^*_{j\|k} \sim N(c^{pop}, \sigma_c);\ \sigma_c \sim N(0,1.0)[0,]$<br>$d^*_{j\|k} \sim N(d^{pop}, \sigma_d);\ \sigma_d \sim N(0,0.5)[0,]$<br>$\hat{e}^*_{j\|k} \sim N(\hat{e}^{pop}, \sigma_{\hat{e}});\ \sigma_{\hat{e}} \sim N(0, 2.0\ )[0,]$ |
| $\sigma_y$ | Standard deviation of the proportion of foci reduction | $\sigma_y \sim N(0, 1)[0,]$ |

[+] Non-centered parameterization was employed for all parameters to improve sampling efficiency. Specifically, for a parameter $\theta \sim N(\mu, \sigma)$, we reparametrized it as $\theta = \mu + \sigma \cdot \tilde{\theta}$, where $\tilde{\theta} \sim N(0, 1)$,

[†] $l$=1,2,3 and 4 indicate a virus working dilution of 1:200, 1:300, 1:330 and 1:400, respectively.

[‡] Parameter bounds are indicated in the form [a, b] for both lower and upper limits, and [a,] when only the lower limit is defined.

**Table S3. Specification of parameter settings for the simulation study.**

| Parameter | Notation | Value |
|---|---|---|
| Mean foci count of VC for each working virus stock | $\delta = [\delta_1, \delta_2, \delta_3, \delta_4]'$ | $[50, 47, 38, 28]'$ |
| 4PL parameters for PC | $\theta^{PC} = [b^{PC}, c^{PC}, d^{PC}, \hat{e}^{PC}]'$ | $[0.945, 0.004, 0.977, 6.524]'$ |
| 4PL parameters for IS500 | $\theta^{IS500} = [b^{IS500}, c^{IS500}, d^{IS500}, \hat{e}^{IS500}]'$ | $[0.984, -0.019, 0.965, 6.357]'$ |
| 4PL parameters for IS1000 | $\theta^{IS1000} = [b^{IS1000}, c^{IS1000}, d^{IS1000}, \hat{e}^{IS1000}]'$ | $[1.068, -0.018, 0.981, 6.808]'$ |
| Population-level mean parameters across all serum samples | $\theta^{pop} = [b^{pop}, c^{pop}, d^{pop}, \hat{e}^{pop}]'$ | $[0.923, -0.028, 0.972, 5.545]'$ |
| **Distribution characteristics of batch-level random effects** ($\boldsymbol{\Gamma} = [\boldsymbol{\gamma_b}, \boldsymbol{\gamma_c}, \boldsymbol{\gamma_d}, \boldsymbol{\gamma_{\hat{e}}}, \boldsymbol{\varsigma}]'$)† | | |
| Mean | $[\mu_{\gamma_b}, \mu_{\gamma_c}, \mu_{\gamma_d}, \mu_{\gamma_{\hat{e}}}, \mu_{\varsigma}]'$ | $[0, 0, 0, 0, 0]'$ |
| Standard deviation | $[\sigma_{\gamma_b}, \sigma_{\gamma_c}, \sigma_{\gamma_d}, \sigma_{\gamma_{\hat{e}}}, \sigma_{\varsigma}]'$ | $[0.028, 0.211, 0.009, 0.250, 3.670]'$ |
| Correlation matrix: | $\rho(\gamma_b, \gamma_c, \gamma_d, \gamma_{\hat{e}}, \varsigma)$ | $\begin{bmatrix} 1 & 0 & 0 & 0 & 0 \\ 0 & 1 & 0 & 0 & -0.7 \\ 0 & 0 & 1 & 0.6 & 0 \\ 0 & 0 & 0.6 & 1 & 0 \\ 0 & -0.7 & 0 & 0 & 1 \end{bmatrix}$ |
| **Distribution characteristics of sample-level random effects** ($\Lambda = [\lambda_b, \lambda_c, \lambda_d, \lambda_{\hat{e}}]'$)‡ | | |
| Mean | $[\mu_{\lambda_b}, \mu_{\lambda_c}, \mu_{\lambda_d}, \mu_{\lambda_{\hat{e}}}]'$ | $[0, 0, 0, 0]'$ |
| Standard deviation | $[\sigma_{\lambda_b}, \sigma_{\lambda_c}, \sigma_{\lambda_d}, \sigma_{\lambda_{\hat{e}}}]'$ | $[0.100, 0.100, 0.006, 1.000]'$ |
| Correlation matrix: | $\rho(\lambda_b, \lambda_c, \lambda_d, \lambda_{\hat{e}})$ | $\begin{bmatrix} 1 & 0 & 0 & 0 \\ 0 & 1 & 0 & 0 \\ 0 & 0 & 1 & 0.6 \\ 0 & 0 & 0.6 & 1 \end{bmatrix}$ |

† Batch-level random effects, collected by a vector $\Gamma = [\gamma_b, \gamma_c, \gamma_d, \gamma_{\hat{e}}, \varsigma]'$, were drawn from a multivariate normal distribution, where $\gamma_b, \gamma_c, \gamma_d, \gamma_{\hat{e}}$ denote effects on each 4PL parameter and $\varsigma$ denote the effect on the mean foci count of VC.

‡ Sample-level random effects, collected by a vector $\Lambda = [\lambda_b, \lambda_c, \lambda_d, \lambda_{\hat{e}}]'$, were drawn from a multivariate normal distribution, where $\lambda_b, \lambda_c, \lambda_d$ and $\lambda_{\hat{e}}$ denote effects on each 4PL parameter.
